# A Novel Sentence Transformer-based Natural Language Processing Approach for Schema Mapping of Electronic Health Records to the OMOP Common Data Model

**DOI:** 10.1101/2024.03.21.24304616

**Authors:** Xinyu Zhou, Lovedeep Singh Dhingra, Arya Aminorroaya, Philip Adejumo, Rohan Khera

**Author notes:** **Correspondence to:** Rohan Khera, MD, MS, 195 Church Street, 6^th^ Floor, New Haven, CT 06510.

## Abstract

Mapping electronic health records (EHR) data to common data models (CDMs) enables the standardization of clinical records, enhancing interoperability and enabling large-scale, multi-centered clinical investigations. Using 2 large publicly available datasets, we developed transformer-based natural language processing models to map medication-related concepts from the EHR at a large and diverse healthcare system to standard concepts in OMOP CDM. We validated the model outputs against standard concepts manually mapped by clinicians. Our best model reached out-of-box accuracies of 96.5% in mapping the 200 most common drugs and 83.0% in mapping 200 random drugs in the EHR. For these tasks, this model outperformed a state-of-the-art large language model (SFR-Embedding-Mistral, 89.5% and 66.5% in accuracy for the two tasks), a widely-used software for schema mapping (Usagi, 90.0% and 70.0% in accuracy), and direct string match (7.5% and 7.5% accuracy). Transformer-based deep learning models outperform existing approaches in the standardized mapping of EHR elements and can facilitate an end-to-end automated EHR transformation pipeline.

## Introduction

Data standards, such as the Observational Medical Outcomes Partnership (OMOP) common data model (CDM), play a crucial role in enabling collaboration across diverse health systems by providing a uniform data standard for organizing EHR (1–5). However, transforming EHR data to the standardized CDMs remains challenging. For instance, a key challenge is the semantic mapping of the EHR elements to their equivalent standard concepts in the CDM. These free-form text elements are often represented in multiple ways in the EHR, limiting the possibility of a one-to-one string matching-based system, which is commonly used in mapping structured elements. Moreover, EHR elements such as drugs present with frequent variations in dosage and frequency, making mapping to the corresponding standardized concepts even more challenging.

Several models have been developed to assist in the matching of EHR elements to CDM concepts, with varying degrees of performance and training requirements. For instance, Usagi is a commonly used software to map the terminologies from EHR to OMOP CDM, based on the term frequency - inverse document frequency (TF-IDF) algorithm (6). Advancements in this field led to the development of Text-based OMOP Knowledge Integration (TOKI) (3). TOKI generates sentence embeddings using deep Recurrent Neural Networks (RNN) and FastText, demonstrating a 10% improvement over Usagi in mapping accuracy (3). However, TOKI’s development relied on 83,000 manually verified mappings, and its performance might not be as good in settings without extensive supervised training data. TOKI was also focused on mapping diagnosis conditions alone(3). There have been no deep learning-based approaches developed explicitly for mapping drug concepts to OMOP CDM in settings without extensive training data. In this study, we sought to develop transformer-based natural language processing models for mapping drug concepts in EHR to OMOP CDM (7). The performance of the mapping systems was applied to map drug concepts within the Yale New Haven Health System to OMOP CDM, and we contrasted its effectiveness with existing mapping approaches.

## Methods

### Data Sources

We obtained concept names (of drugs, and all other domains, such as condition, procedure) (n=9,217,224) for model pre-training and their mappings relations (n=4,569,103) for model finetuning, from the Observational Health Data Sciences and Informatics (OHDSI) Vocabularies, accessed through Athena, a publicly available online repository for medical vocabularies (8). These mappings pair a non-standard concept or synonym with a standard concept (Figure 1). Non-standard concepts are concepts in a non-standard code system, where non-standard-to-standard-mappings associates them into the ones in a standard code system. Synonyms, on the other hand, do not exist in code systems, and are alternative names or descriptions for concepts. Standard concepts refer to unified, normalized representations of medical terminologies for organizing and standardizing healthcare data. For instance, both the non-standard concept “IRON 325 MG TABLET” and synonym “FESO4 325 MG Oral Tablet” can be mapped to the standard concept “ferrous sulfate 325 MG Oral Tablet”. To pre-train models in a self-supervised style, we collected all unique concept names and concept synonym names from Athena vocabulary.

**Figure 1.**
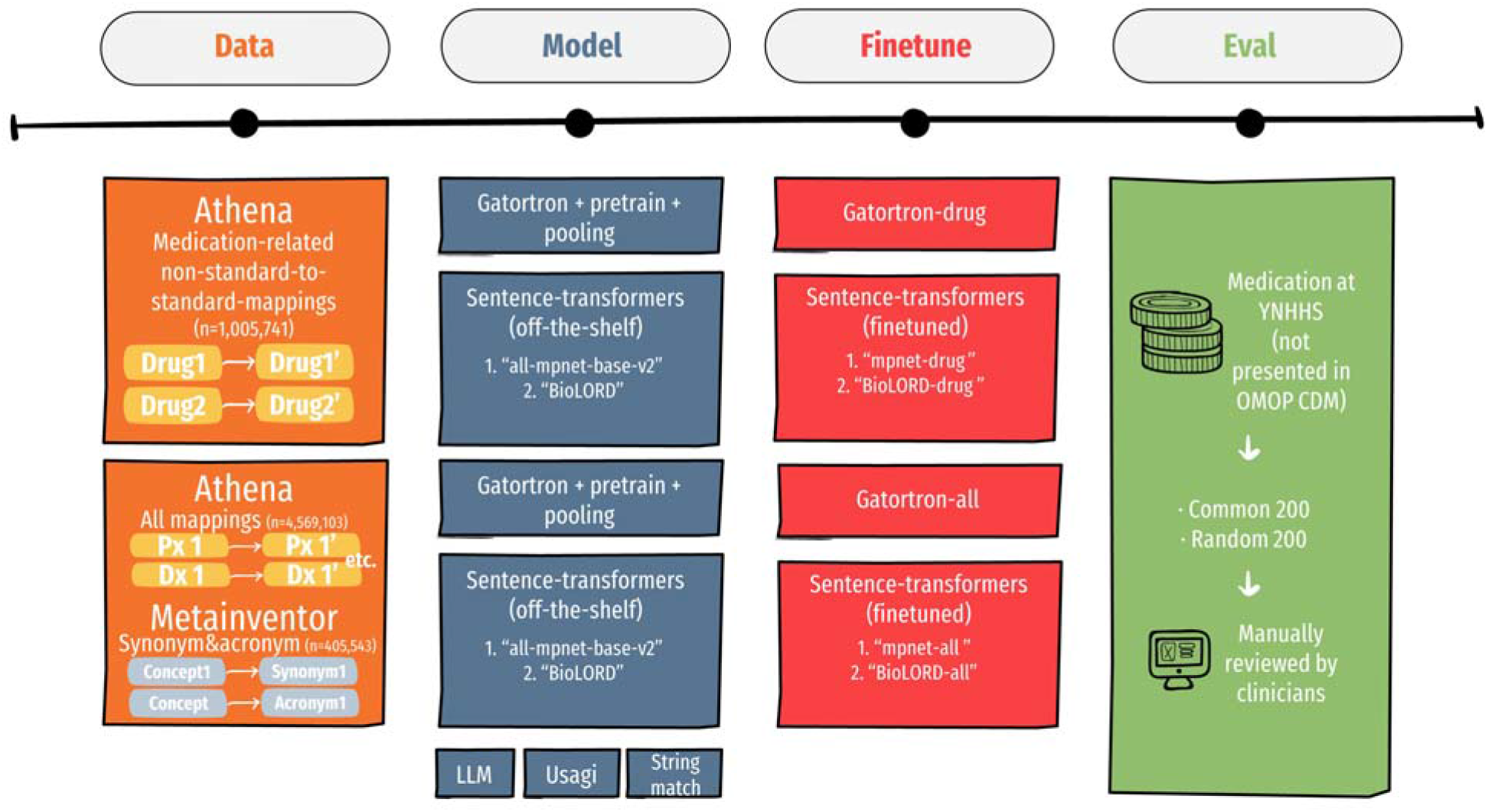
An overview of this study. Using data from the Athena vocabularies and mapping relationships on Metainventor medical acronyms and abbreviations database, we pretrained and finetuned off-the-shelf models. The clinician manually evaluated a total of 11 approaches to map medication concepts in YHNNS’s EHR to OMOP CDM.

Additionally, we assembled medical acronyms and abbreviations from the Metainventor database for model finetuning (n=405,543).(9) Each record in Metainventor is also a mapping pair, where a medical concept is mapped to its acronym(s) or abbreviation(s). (Figure 1)

To evaluate the effectiveness of the mapping approaches, we collected the drug concept names from the structured medication table from a cohort drawn from the EHR at YNHHS. YNHHS is the largest healthcare network in Connecticut, comprising five hospitals and a broad outpatient provider system. All unique drug concept names were sourced from the Clarity database, a comprehensive SQL-based reporting tool from Epic Systems Corporation, extracting data from the YNHHS EHR system’s medication table.

### Model Development

We followed the sentence-transformer approach to develop our models. (10, 11) Sentence-transformers typically consist of a pre-trained transformer *encoder* with an overlaying pooling layer. A drug concept is consisting of one or multiple tokens *t*_1_, *t*_2_, …, *t*_n_, where each token is a sub-word. The *encoder* generate an embedding (i.e., high dimensional vectors) *E*_1_, *E*_2_, …, *E*_n_ for each token *t*_1_, *t*_2_, …, *t*_n_. All sentence-transformer models in this study utilized a mean pooling layer to generate *E*, a unified embedding for a drug concept, based on all token-level embeddings of the drug concept: 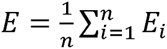. There are publicly available sentence-transformer models, which are commonly already trained with sentence pairs to produce meaningful sentence-level embeddings. The model was trained to maximize the distance between embeddings of dissimilar concepts and minimize the distances between similar concepts. (10, 11) We trained models in a similar style, and clinicians in our team evaluated both off-the-shelf models and the models we trained.

In assessing readily available sentence-transformer models, we evaluated the accuracy of an off-the-shelf leading general-purpose sentence-transformer (all-mpnet-base-v2) alongside a premier off-shelf clinical sentence-transformer (BioLORD) (10, 11).

To improve the embeddings generated by sentence-transformers and facilitate accurate clinical terminology mapping, we trained sentence-transformer models using publicly available clinical mapping relationships, yielding six models: (3) mpnet-drug (4) BioLORD-drug (5) mpnet-all (6) BioLORD-all (7) Gatortron-drug (8) Gatortron-all. The models were trained with multiple negatives ranking loss. The loss function minimized the distance between the embedding of mapping pairs, while maximizing the distance between the embedding of negative pairs. Within each batch containing N mapping pairs (a_1_, p_1_), …, (a_n_, p_n_) for a mapping pair (a_i_, p_i_), the negative pairs are all N-1 (a_i_, p_k_) where i ≠ k. MultipleNegativesRankingLoss can be expressed as follows:

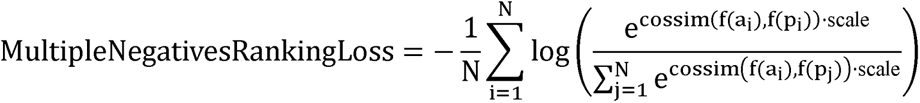

where f(·) is the transformer-based natural language processing model which turns a medication terminology into an embedding. We used cosine similarity (cossim) to calculate the distances between embeddings. scale is a hyperparameter for changing the sensitivity of MultipleNegativesRankingLoss towards inaccurate embeddings, and we used its default value of 20.

By training the two aforementioned sentence-transformer models (1) all-mpnet-base-v2 (2) BioLORD, respectively, using drug non-standard-to-standard-mappings (from Athena Vocabularies, with 1,005,741 training pairs), we created drug mapping models (3) net-drug (4) BioLORD-drug. We also trained the off-the-shelf sentence-transformer models (1) all-mpnet-base-v2 (2) BioLORD using our full supervised training set consisting of 4,569,103 mapping pairs (including both non-standard-to-standard-mappings and synonym relationships) from Athena Vocabularies and 405,543 medical acronyms and abbreviations from Metainventor. Such training resulted in two additional models: (5) mpnet-all and (6) BioLORD-all. These models were trained for 10 epochs, with a batch size of 96.

We also developed two sentence-transformer models based on a BERT-like model by a) self-supervised pretraining and b) supervised training using mapping pairs. GatorTron-Base is a encoder-only public-available model with 345 million parameters pre-trained using deidentified clinical notes at the University of Florida (12). During the a) self-supervised pretraining step, we pursued continual pre-trained GatorTron-Base via masked language model objective using drug-related concept names and synonyms (n=5,185,133 terminologies), or all concept names from OHDSI (n=9,217,224 terminologies) (12–14), yielding two encoder-only models. We continually pre-trained the models for 3 epochs, with a maximum input length of 64 and a default masking probability of 15%. We added a mean pooling layer to the continual pre-trained encoder-only models. These models can generate a respective embedding for each input. During the b) supervised training using mapping pairs phrase, we trained models with mapping pairs using a batch size of 48 for 10 epochs. The model continually pre-trained using drug vocabulary was trained with drug non-standard-to-standard-mappings (n=1,005,741 training pairs from Athena Vocabularies), and it’s called (7) Gatortron-drug. For the model continual pre-trained with all vocabularies, we trained it with all mapping pairs from Athena Vocabularies (n=4,569,103 mapping pairs) and medical acronyms and abbreviations from Metainventor (n=405,543), yielding a model called (8) Gatortron-all.

We developed our models fully based on publicly available data and evaluated them on Yale’s EHR data in a secure environment without further training to demonstrate that the approaches developed may be applied in other healthcare systems as well.

### Mapping to OMOP CDM

Sentence-transformer models can convert each drug concept name (i.e., terminology) into one high-dimensional vector (i.e., embedding). Cosine similarities were calculated between each embedding of terminology at YNHHS and embeddings of all standard drug concepts in OMOP CDM. The best mapping was the one that maximizes the cosine similarity (7, 15).

Similarly, we evaluate the mapping outputs of an embedding approach using a large language model (LLM) (SFR-Embedding-Mistral) with over 7 billion parameters and state-of-the-art performance in the Massive Text Embedding Benchmark (MTEB) benchmark (17, 18), a commonly used software (Usagi) for clinical concept mapping, and a string match approach (Python package RapidFuzz) as the baseline. SFR-Embedding-Mistral is an LLM based on Mistral 7B (16, 17). It appends a [EOS] token to the end of the input before feeding it to the LLM (17). The embedding was the hidden vector in the last layer corresponding to the [EOS] token, which has been finetuned using large-scale sentence pair datasets. Usagi is based on the TF-IDF algorithm (15). TF-IDF evaluates the similarity of medical terminologies by paying attention to the words with low occurrence in all terminologies (such as “Ibuprofen”) rather than common words (such as “gram”). Meanwhile, RapidFuzz converts drug concept names to token sets and computes the Levenshtein Distances between token sets to find the optimal mappings.

### Statistical Analysis

We identified the 200 most common medications given to the patients at YNHHS and 200 random medications in the YNHHS EHR database (excluding those not in RxNorm format, which are the standard OMOP concept) and aligned them with standard concepts in the OMOP CDM. The outputs of the models were evaluated by clinicians (LSD and AA) independently. We report the number of model errors, distinguishing between incorrect ingredient identification and correct ingredient but incorrect dosage. We presented model accuracies alongside their confidence interval calculated using Python package “statsmodels”. Chi-squared tests were employed to detect if the differences in model performances were statistically significant (p<0.05). All statistical analyses were performed using Python 3.9.

## Results

### Study Population

We used data from a cohort of 146,397 patients at the YNHHS. Across 12,543,715 rows of data in the medication dataset, there were 39,441 unique medications – 36,212 (92%) of which were not present in RxNorm – the standard medication code system in OMOP CDM. The most frequently prescribed 200 medications constituted 3,885,163 (31.0%) of all medication orders.

### Model Performance Across Most Common Medications

We collected the 200 most common drug concepts that are not presented in OMOP CDM. Eleven approaches were deployed to map the drug concepts at YNHHS to OMOP CDM (Table 1). Usagi (a commonly used software based on TF-IDF) reached 90.0% accuracy, while SFR-Embedding-Mistral (the state-of-the-art off-the-shelf LLM) reached an accuracy of 89.5%. Among off-the-shelf sentence-transformers, BioLORD, a clinical sentence-transformer, displayed an accuracy of 92.0%. Meanwhile, all-mpnet-base-v2, one of the best general-purpose sentence-transformers, displayed a lower accuracy of 62.0%. String match-based mapping yielded a low accuracy at 7.5%.

**Table 1.** Comparison of model architecture, pretraining and training data sources, and error statistics in mapping the 200 most common drug concepts.

| Approach or Model | Algorithm or backbone model | Number of parameters | Data sources for additional domain-specific model pre-training and training | Errors on the ingredient | Errors on the dosage | Total errors | Accuracy (95% CI) |
| --- | --- | --- | --- | --- | --- | --- | --- |
| RapidFuzz | String match, bag-of-words | 0 |  | 176 (88.0%) | 9 (4.5%) | 185 (92.5%) | 7.5% [3.8% - 11.2%] |
| Usagi | TF-IDF, bag-of-words | 0 |  | 7 (3.5%) | 13 (6.5%) | 20 (10.0%) | 90.0% [85.8%, 94.2%] |
| all-mpnet-base-v2 |  | 133M |  | 4 (2.0%) | 72 (36.0%) | 76 (38.0%) | 62.0% [55.3% - 68.7%] |
| BioLORD |  | 133M |  | 2 (1.0%) | 14 (7.0%) | 16 (8.0%) | 92.0% [88.2% - 95.8%] |
| SFR-Embedding-Mistral |  | 7.11B |  | **0 (0.0%)** | 21 (10.5%) | 21 (10.5%) | 89.5% [85.3%, 93.7%] |
| mpnet-drug | all-mpnet-base-v2 | 133M | Pre-training: NA<br>Training: drug mappings from Athena Vocabulary | 2 (1.0%) | **5 (2.5%)** | **7 (3.5%)** | **96.5% [94.0% - 99.0%]** |
| BioLORD-drug | BioLORD | 133M |  | 2 (1.0%) | 8 (4.0%) | 10 (5.0%) | 95.0% [92.0% - 98.0%] |
| Gatortron-drug | GatorTron-base | 345M | Pre-training: drug concept and concept synonyms from OHDSI vocabulary<br>Training: drug mappings | 1 (0.5%) | 9 (4.5%) | 10 (5.0%) | 95.0% [92.0% - 98.0%] |
| mpnet-all | all-mpnet-base-v2 | 133M | Pre-training: NA<br>Training: all mapping pairs from Athena Vocabulary | 13 (6.5%) | 13 (6.5%) | 26 (13.0%) | 87.0% [82.3% - 91.7%] |
| BioLORD-all | BioLORD | 133M |  | 13 (6.5%) | 19 (9.5%) | 32 (16.0%) | 84.0% [78.9% - 89.1%] |
| Gatortron-all | GatorTron-base | 345M | Pre-training: all concepts from Athena Vocabulary<br>Training: all mapping pairs from Athena Vocabulary | 16 (8.0%) | 26 (13.0%) | 42 (21.0%) | 79.0% [73.4% - 84.6%] |

CI: 95% confidence interval

When trained with drug mapping collected from OHDSI vocabularies, all three transformer-based models outperformed SFR-Embedding-Mistral (the LLM) and Usagi (the commonly used software), reaching accuracies ≥95.0%. In particular, mpnet-drug reached the highest accuracy at 96.5%. It outperformed the state-of-the-art LLM-based embedding approach (p=0.011), the software based on TF-IDF (p=0.017), and the best off-the-shelf clinical sentence-transformer (p=0.086). Compared to the off-the-shelf approaches, the mpnet-drug model was more accurate about the ingredient and dosage of drugs when mapping. However, after training with our full training set, which contains mapping relations other than medication, synonyms, acronyms, and abbreviations, the performance of transformer-based deep learning models did not outperform Usagi and LLM.

### Model Performance Across a Random Subset of Medications

Models were applied to map a random sample of 200 unique drug concepts in YNHHS’s EHR that were not present in OMOP CDM, and model performances were evaluated (Table 2). Among off-the-shelf approaches, BioLORD (a clinical sentence-transformer, accuracy: 71.5%), Usagi (a commonly used software based on TF-IDF, accuracy: 70.0%), and SFR-Embedding-Mistral (the state-of-the-art off-the-shelf LLM, accuracy: 66.5%) reached relatively high performance. Meanwhile, all-mpnet-base-v2, one of the best off-the-shelf sentence-transformer models for general settings, only reached 48.0% accuracy. The string matching had the same accuracy for these sets of drugs as the most commonly used ones, at 7.5%.

**Table 2.**
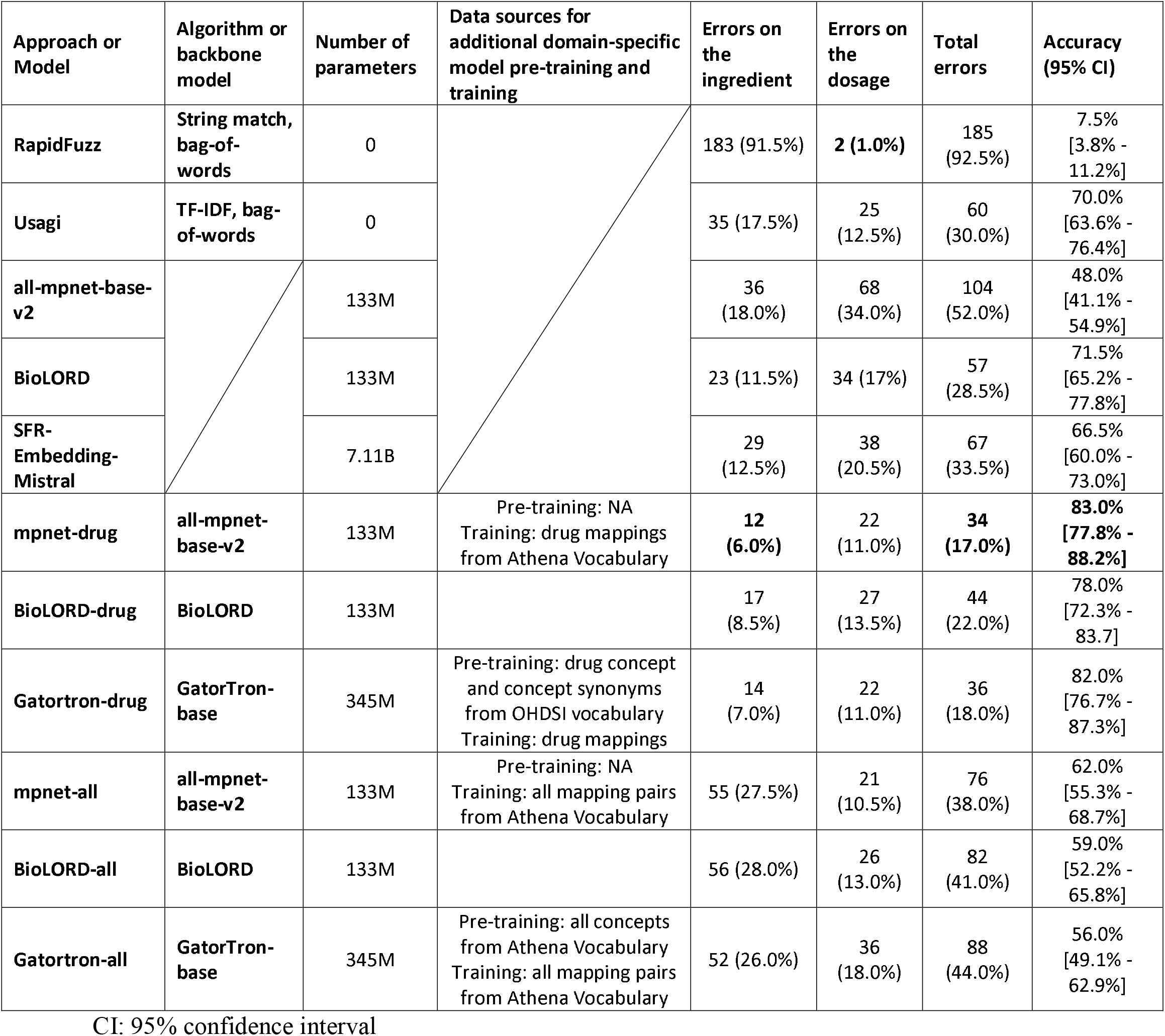
Comparison of model architecture, pretraining and training data sources, and error statistics in 200 random drug concepts across models.

After training with drug mappings collected from OHDSI vocabularies, all transformer-based deep learning approaches reach higher accuracies than the off-the-shelf approaches, with reduced error both in the ingredient and dosage of drugs when mapping. mpnet-drug, a model with all-mpnet-base-v2 as its backbone model, reached the highest accuracy (83.0%). It outperformed the best off-the-shelf clinical sentence-transformer (p=0.009), Usagi (p=0.003), and a state-of-the-art LLM (p<0.001). In addition, Gatortron-drug reached 82.0% accuracy, and BioLORD-drug reached 78.0%, both higher than the off-the-shelf approaches. Still, after training with the full complete training set (which contained mapping relations other than medications, as well as acronyms and abbreviations), the transformer-based deep learning models did not reach higher performance than the best off-the-shelf approach.

## Discussion

In this study, we trained transformer-based natural language process models on publicly available datasets to enable the mapping of drug records at YNHHS to the OMOP CDM without need development on protected clinical EHR data. Our top-performing model achieved state-of-the-art accuracies in mapping most common medications and a random subset of medication given to the patients, significantly exceeding both Usagi (a commonly used software based on TF-IDF) and SFR-Embedding-Mistral (the state-of-the-art off-the-shelf LLM), with fewer errors due to both the dosages and ingredients.

In this regard, our approach achieved the benchmark of outperforming Usagi met by TOKI, a previously developed supervised deep learning-based approach (3). Compared with TOKI – which was built on traditional deep learning techniques, including RNN and FastText, our approach incorporates recent progress in deep learning, including embeddings from pretrained transformers encoders (7, 13). We further leveraged masked language model pretraining and trained the models with millions of sentence pairs to boost the model performance. Leveraging superior model architecture and large-scale publicly-available datasets enables state-of-the-art accuracy without training on YNHHS’s data. Thus, the time-consuming and expensive annotation of supervised datasets is not needed before applying our approach to map private EHR to OMOP CDM. Of note, TOKI was evaluated only in condition mapping, whereas our model was developed to focus on drug records mapping, a key operational priority and a more complex task. Since the Athena vocabularies also contain mapping relationships for all other domains, spanning conditions, procedures, and measurements. In this way, our transformer-based approach might be expandable to additional domains (8), and similar models can be developed to facilitate automated end-to-end EHR to OMOP CDM to facilitate generalizability.

In settings where automated mapping system to OMOP CDM is needed, a high-quality supervised training set, like the one for developing TOKI (3), may not always be readily available. Our approach was developed using publicly available data and evaluated on protected EHR at YNHHS. The models are robust despite the vocabularies only reporting some key relationships and no site-specific training names of the medications recorded within the EHR at Yale. We anticipate an increase in performance if further site-specific training on the distribution of words is sought, but by ensureing the models were not trained in YNHHS data, our approach would be more likely to generalize to other hospital systems without need for local development. Our study has certain limitations. We only evaluated the models on drug concepts at YNHHS. Therefore, our approach’s effectiveness in other domains (like conditions and procedures) and at other hospital systems remains untested. However, conditions and procedures are often readily mapped using standard ontologies like ICD or CPT to SNOMED mapping. Also, our approach can be applied to map other domains, thanks to the availability of mapping pairs of other domains on Athena Vocabulary. Another limitation is we did not leverage LLMs with better performance, such as ChatGPT, into the workflow. Future studies may explore finetuning ChatGPT with the dataset described in this study to develop a reliable end-to-end mapping pipeline without human in the loop, and therefore aligning EHR in many hospital systems at a relatively low cost.

## Conclusion

Sentence transformer-based natural language processing models can enable automated mapping concepts of drugs at YNHHS to counterparts in OMOP CDM automatically with significantly improved accuracy. Similar approaches can be applied in other domains and organizations (5).

## Data Availability

The model was evaluated on protected health information, which cannot be shared publicly to protect patient confidentiality. The OMOP dictionaries used to train the model are available at:
https://athena.ohdsi.org/vocabulary/list

